# Modelling vaccination and control strategies of outbreaks of monkeypox at gatherings

**DOI:** 10.1101/2022.08.12.22278724

**Authors:** Pei Yuan, Yi Tan, Liu Yang, Elena Aruffo, Nicholas H. Ogden, Jacques Bélair, Julien Arino, Jane Heffernan, James Watmough, Hélène Carabin, Huaiping Zhu

**Affiliations:** Laboratory of Mathematical Parallel Systems (LAMPS), Department of Mathematics and Statistics, York University, Toronto, Canada; Canadian Centre for Diseases Modeling (CCDM), York University, Toronto, Canada; School of Mathematics and Statistics, Northeast Normal University, Changchun, Jilin, China; Public Health Risk Sciences Division, National Microbiology Laboratory, Public Health Agency of Canada; Département de Mathématiques et de Statistique, Université de Montréal, Montréal, Québec, Canada; Department of Mathematics, University of Manitoba, Winnipeg, Manitoba, Canada; Department of Mathematics and Statistics, York University, Toronto, Canada; Department of Mathematics and Statistics, University of New Brunswick, Fredericton, New Brunswick, Canada; Département de Pathologie et Microbiologie, Faculté de médecine vétérinaire, Université de Montréal, Saint-Hyacinthe, Québec, Canada; Département de médecine sociale et préventive, École de santé publique de l’université de Montréal, Montréal, Québec, Canada; Centre de Recherche en Santé Publique (CReSP) de l’université de Montréal et du CIUSS du Centre Sud de Montréal, Montréal, Québec, Canada; Groupe de Recherche en Épidémiologie des Zoonoses et Santé Publique (GREZOSP), Université de Montréal, Saint-Hyacinthe, Québec, Canada

**Keywords:** Monkeypox, vaccination strategy, ring vaccination, gatherings, testing, modelling, control

## Abstract

**Background:** Monkeypox cases keep soaring in non-endemic’s countries and areas in the last few months, leading to the WHO declaring a Public Health Emergency of International Concern. The ongoing and coming festivals, parties and holidays gathering events are causing increased concerns about possible outbreaks.

**Methods:** We considered a hypothetical metropolitan city and modelled the transmission of monkeypox virus in humans in high-risk (HRG) and low-risk groups (LRG) using a Susceptible-Exposed-Infectious-Recovered (SEIR) model and incorporated gathering events. Model simulations assessed how the current vaccination strategy combined with other public health measures can contribute to mitigating or halting outbreaks from mass gathering events.

**Results:** The risk of a monkeypox outbreak remains high on the occasion of mass gathering events in the absence of public health control measures. However, the outbreaks can be well controlled by cutting off transmission by isolating confirmed cases and inoculating their close contacts. Also, Post Exposure Prophylaxis is more effective for containment in the summer gatherings than a broad vaccination campaign in HRG, considering the time needed for developing the immune response and the availability of vaccine. The number of attendees and effective contacts during the gathering are the factors that need more attention by public health authorities to prevent a burgeoning outbreak. Moreover, restricting attendance through vaccination requirements can help secure mass gathering events.

**Conclusion:** Gathering events can be made safe with some restrictions of either the number and density of attendees in the gathering, or vaccination requirements. The ring vaccination strategy inoculating close contacts of confirmed cases may not be enough to prevent potential outbreaks, however, mass gatherings can be rendered safe if that strategy is combined with public health measures, including rigorous contact tracing, testing, identifying and isolating cases. Compliance of the community and promotion of awareness are also indispensable to the containment.

## 1. Introduction

Monkeypox, a zoonosis, has been recorded since early May 2022 in at least 30 non-endemic countries such as Spain, the United States, Germany, the United Kingdom, France, Canada^1^. As of July 21, 2022, the cumulative number of confirmed cases exceed 15,000 globally^2^. On July 23, 2022, the World Health Organization (WHO) declared monkeypox a Public Health Emergency of International Concern (PHEIC) due to outbreaks in multiple countries and continents^3^. The Public Health Agency of Canada (PHAC) reported 539 confirmed cases of monkeypox by July 15, 2022, including 32 confirmed cases in British Columbia, 12 in Alberta, 2 in Saskatchewan, 194 in Ontario, and 299 in Québec^4^. The unusual outbreaks emerged in non-endemic areas and the associated increasing number of cases raise serious concerns about the spread within communities and areas, and undetected cases, especially among high-risk populations such as health-care providers, gay, bisexual and other men who have sex with men (gbMSM)^5^. Understanding the spreading mechanism and seeking potential control of the current monkeypox outbreak are thus of pressing wide interest^6^.

At present, PHAC in Canada, where all authors are based, has adopted measures such as vaccines, testing, tracking and isolation to closely monitor the course of infection and transmission risk of the disease in Canada^7^. With the increasing threat of monkeypox in the country, PHAC has been working with provincial and territorial public health partners to closely monitor, detect and confirm cases. However, Canada is likely weeks behind in identifying the true scope of monkeypox spread due to the limited testing resources. On the other hand, vaccines have been proven extremely useful to help eradicate and prevent many infectious diseases^8,9^. As a successful example of pharmaceutical measures, smallpox vaccines were used during the global smallpox eradication programs^10^. Hence, the smallpox vaccines are administered to contain the current monkeypox spreading in Canada. In June 2022, the National Advisory Committee on Immunization (NACI) released a guideline on using an orthopoxvirus (Imvamune®) vaccine with potential efficacy against monkeypox^11^. The guideline recommends vaccination for adults at high risk of occupational exposure for Pre-Exposure Prophylaxis (PrEP) and Post-Exposure Prophylaxis (PEP). In May 2022, PHAC placed an order for 500,000 vials of the smallpox vaccine Imvamune. However, these doses will not be delivered until April 2023^12^, therefore the mass vaccination campaign in the broad population is not possible given the limited stockpiles of vaccines. Hence, the appropriate vaccination rollout strategy is much needed for public health.

After two years of restrictions on gatherings due to the control and prevention of COVID-19 epidemics, mass gatherings for festivals and ceremonies are allowed with no attendance limitations^13^. In many Canadian provinces, local festivals recorded attendance close to the pre-pandemic leve^14-16^ which has led to some concerns on the spread and possible outbreaks of monkeypox. WHO also expressed concerns that more infections could arise in Europe and elsewhere^17^ due to private and social gatherings during festivals, parties and holidays. In fact, in the United States, many cases were reported linked to large social gatherings, such as Pride events, or pool parties and bathhouses^18,19^. Consequently, it is essential to assess the effect of gathering events on disease transmission to inform public health for potential implementations of control measures.

Transmission risk at a gathering is mainly associated with the gathering size^20^ and is proportional to the population density at the gathering place^21^. Using an individual-based model, Liu et al.^22^ showed that for mass gathering event (MGE) with 200,000 participants, there is a 23.6% increase of positive cases attributed to MGE for the transmission of COVID-19. The effect of increased density of contacts during Hajj was estimated to generate a 78-fold increase in disease transmission that impacts not only pilgrims but also the local population^23^.

To investigate the dynamics of monkeypox and provide information to public health for prevention and control, especially at gatherings, we establish a Susceptible-Exposed-Infectious-Recovered (SEIR) modelling framework with a prodromal stage to assess the effect of the current vaccination strategy. The PrEP and PEP, testing and isolation, as well as the intervention of gathering events are also considered. We mainly focus on assessing the effectiveness of vaccination and public health control measures to simulate the scenarios of gathering with different numbers of attendees and different levels of interventions to inform public health decision-making. Our findings confirm that the ring vaccination approach itself is not enough; however, inoculating the close contacts of identified cases can be effective to prevent the burgeoning outbreak after the mass gathering events, which contributes to cutting off the routes for further spreading, if followed with timely testing and compliance with the isolation.

## 2. Methods

### 2.1 Modelling overview

The vast majority of current cases are linked to specific high-risk locations and populations. Hence, to better capture the infection dynamics within different risk settings, we consider the population to be divided into two subgroups: low-risk population (LRG), which defines the individuals who behave to reduce the possibility to become infected, and the high-risk population (HRG), which lives and behaves with a higher chance of becoming infected. For simplicity, henceforth we use the subscript 1 for the LRG, and 2 for the HGR. Those two groups interact between and within groups as represented by a contact matrix (*c*_*ij*_, *i,j* = 1,2), defined using the assumptions in Yuan et al^24^. We assume that there is no movement of population between the risk groups, unless there is a gathering event such that a proportion of LRG people may become part of the HRG.

The infection dynamic follows the Susceptible-Exposed-Infectious-Recovered (SEIR) framework, which is extended to include prodromal stage, vaccination, testing and isolation. Susceptible individuals become infected, and move to the exposed compartment, after encountering an infectious individual from either LRG and HGR, assuming that the latter group is more infectious than the first group. After a latent period, the prodromal stage begins, and during this phase no symptoms are apparent, but the individual can shed the virus. This period is then followed by the infectious stage and then recovery occurs. Infectious individuals with symptoms may be tested, then confirmed and isolated, while their contacts, which might be susceptible, exposed, or pre-symptomatic, can be vaccinated and quarantined to prevent any further spread of the infection. Although the model does not include demographics, we assume infection-related death might occur among infectious individuals in both risk groups. Isolated individuals who develop infection will remain isolated until recovery. Individuals in both prodromal and infectious stages can transmit the infection, however LRG people are assumed to be less infectious than those in the HRG. The population structure and flow diagram of the disease are shown in **Figure 1**.

**Figure 1.**
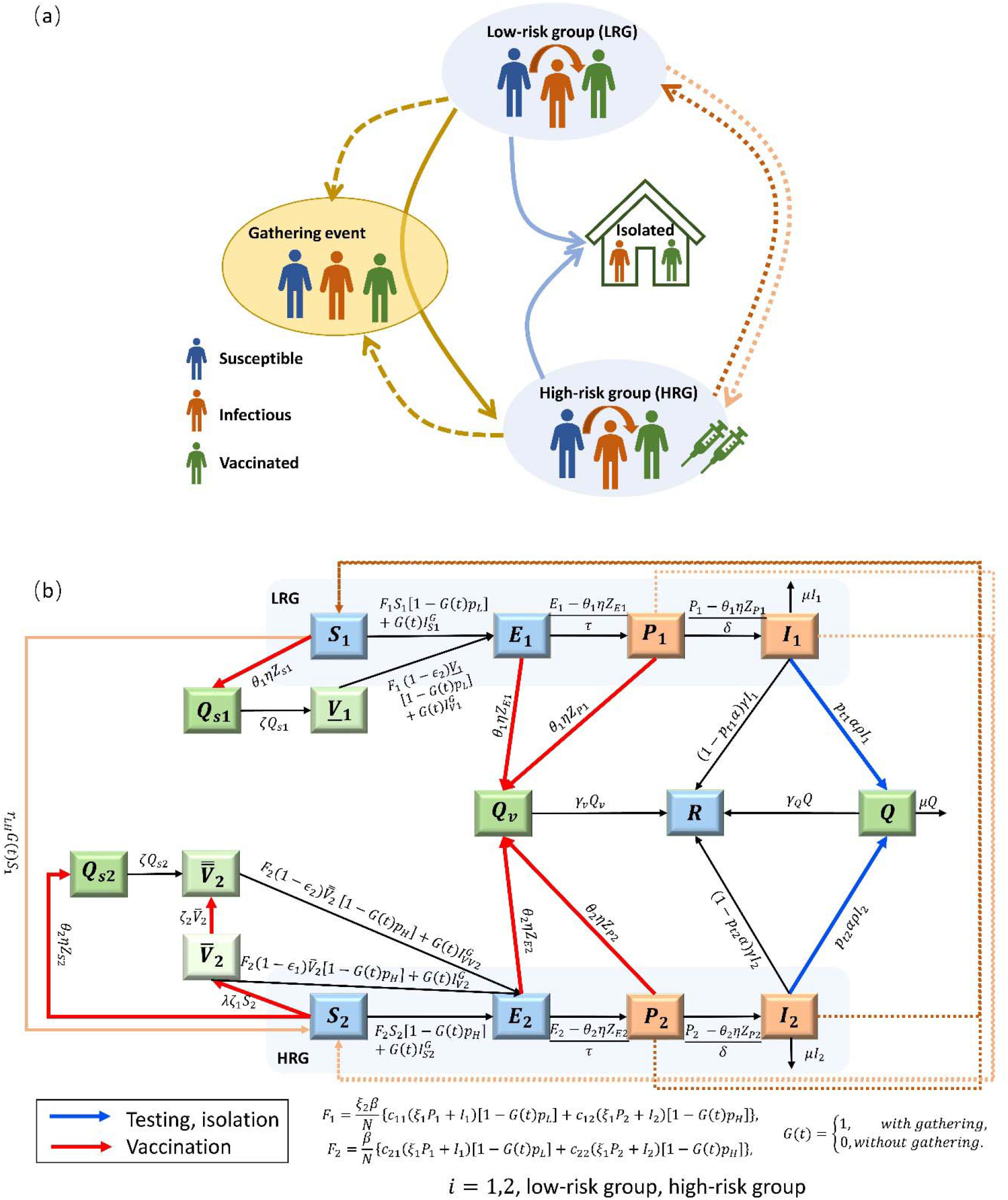
Schematic diagram (a) and flow chart (b) of the MPX transmission among the population classified with low-risk group (LRG, i=1) and high-risk group (HRG, i=2), considering the gathering event.

We include the vaccination process announced by PHAC^11^ to explore its effectiveness by assuming that only high-risk people will be vaccinated. Given the extensive vaccine campaign against smallpox until 1972, we assume that all individuals born before that year are fully vaccinated therefore protected. Except for the vaccination, we also examine the public health measures of testing and isolating. Model assumptions, variables and parameters are summarized in Tables 1-3, and the model equations are presented in **Appendix A1**.

**Table 1.**
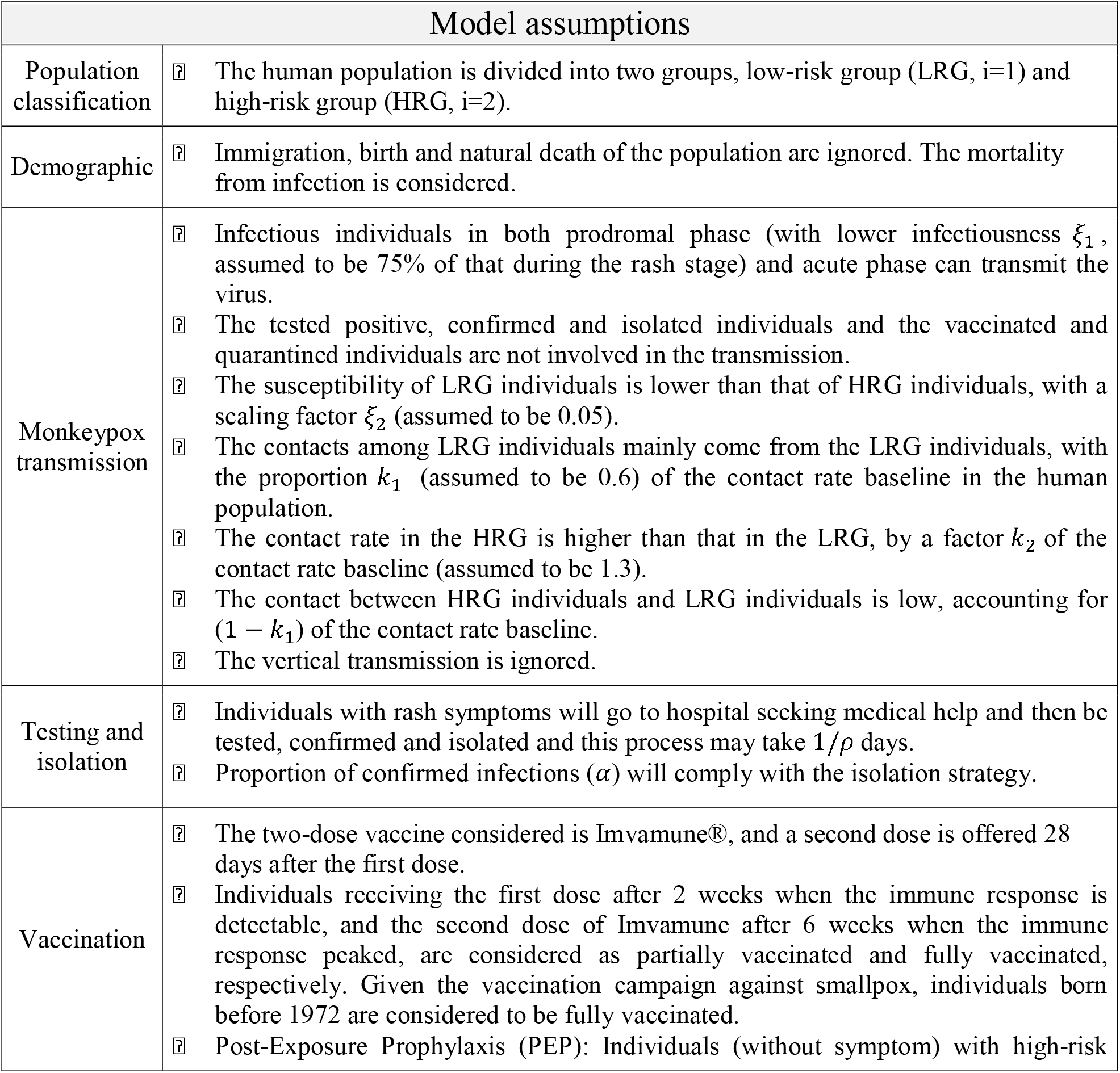
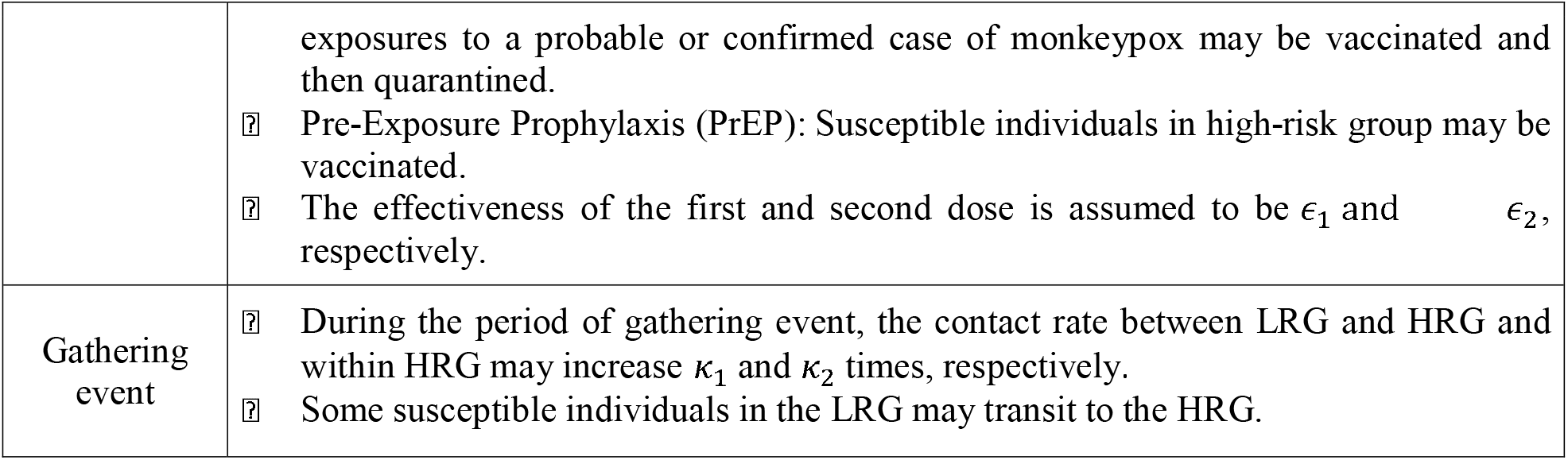
Model assumptions.

### 2.2 Transmission

We use the assumptions of Yuan et al^24^ on the transmission of the monkeypox virus. The probability of transmission per contact was assumed to be 12.2% to 24.5%, and between 0.37% and 0.74%, respectively, among HRG and LRG, as calculated from the basic reproduction number *R*_0_ derived from our simplified model without public health control measures (see **Appendix A2** for details) all the other parameters being fixed (**Table 3**).

### 2.3 Vaccination

On June 10, 2022, NACI issued Interim guidance on the use of Imvamune® in the context of monkeypox outbreaks in Canada^11^. Imvamune, initially developed for the prevention of smallpox, is a two-dose vaccine with the second dose administered 28 days after the first one. The immune response is detectable by week 2 after the first dose and peaks at week 6 after dose 2 in a randomized, open-label trial designed to compare the effectiveness of Imvamune with the second-generation replicating smallpox vaccine^11^. Given the recent emergence of cases and use of vaccines, there is no available data indicating the effectiveness of Imvamune vaccination against monkeypox infection; however, studies of vaccine effectiveness (VE) of smallpox vaccine may provide a general estimation. In the context of PEP, the median effectiveness in preventing smallpox disease with vaccination at 1–3 days after exposure was estimated as 80%^25^. There are too many uncertainties about the effectiveness of Imvamune against monkeypox infection, although some observational studies suggest that the vaccine against smallpox may be about 85% effective in preventing monkeypox^11^. Hence, in our model, we assume that the effectiveness of the first and second doses ranges between 40 and 60%, and 70% and85%, respectively.

#### Post-Exposure Prophylaxis (PEP)

Individuals (without symptoms) with exposure to any confirmed case of monkeypox may be vaccinated and then quarantined, our model will incorporate the monkeypox vaccine PEP for protecting individuals with exposure to confirmed cases. Note that the effectiveness of the vaccine in PEP is not considered since these individuals with exposure will be quarantined and not induce further transmission. Vaccinated and quarantined individuals, with exposure to confirmed cases, will recover during the quarantine if the vaccine is not effective and will not transmit the virus to others.

We denote by 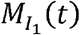 and 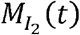 the daily new confirmed monkeypox cases in the LRG and HRG at time t, respectively, and they are defined as:

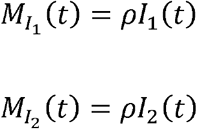

where 1/*ρ* is the average number of days infectious individuals spend between showing rash symptoms to being tested and confirmed.

Following Yuan et al^24^, the number of close contacts of newly confirmed cases in the LRG, who are in exposed state Z_*E*1_(*t*) and prodromal phase *Z*_*P*1_(*t*) at time *t* can be calculated as

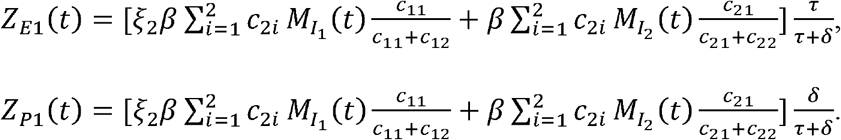

Similarly, we obtain the number of close contacts of newly confirmed cases in the HRG, who are in exposed state *Z*_*E*2_(*t*) and prodromal phase *Z*_*P*2_(*t*), as

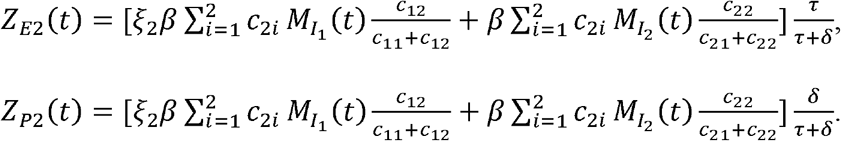

Also, the number of close contacts of newly confirmed cases that are susceptible in the LRG and HRG is

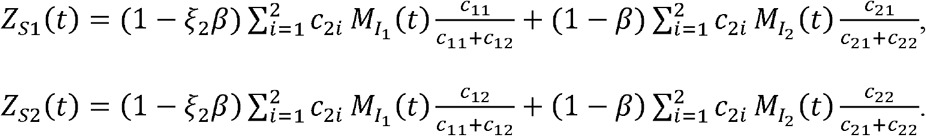

#### Pre-Exposure Prophylaxis (PrEP)

Monkeypox vaccine PrEP refers to administering vaccine to individuals at high risk of exposure to the virus. Hence, in our model, vaccination is only administered to those in HRG. Since people born before 1972 have been vaccinated with the smallpox vaccine, therefore, in the initial state in the model, a proportion of the LRG and HRG populations is considered fully vaccinated for simplicity.

### 2.4 Testing and isolation

Testing and isolation are crucial steps to detect monkeypox infections and stop the virus from spreading. In Canada, the diagnostic tests for monkeypox cases are performed in the Canada’s National Microbiology Laboratory^26^. The turnaround time of testing varies among the Canadian provinces, and it ranges from 36 to 48 hours^27,28^. Individuals with clinical illnesses where monkeypox is suspected should be tested and conduct self-isolation before the negative test result is received, while individuals with positive test results should isolate at home until they recover. However, it can take several days from when infected individuals develop symptoms to seek medical help and then get tested, with large variations depending on the individuals’ behaviours. In addition, the recovery period takes 2 to 4 weeks, and the respect of the isolation strategy may not be total, hence compliance with the isolation strategy for those tested and confirmed monkeypox cases are also included in our model.

### 2.5 Gatherings

Gatherings may contribute significantly to the spread of infectious diseases, as was extensively studied during the SARS-CoV-2 pandemic^20^. Not only will individuals from HRG attend the gathering event, but also individuals in the LRG will join and some of them may transit to the HRG, therefore facilitating virus spreading and posing the risk of a possible outbreak. In our model, we denote *G* (*t*) as an indicator parameter if there is a gathering event.

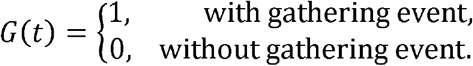

The daily transition rate from the low-risk susceptible individuals to the high-risk is *r*_*LH*_ days^-1^. During the period of gathering events, we assume that the total number of attendees is *N*_*G*_ and the proportion of attendees from LRG and HRG is *p*_*GL*_ and *p*_*GH*_, respectively. The proportion of LRG and HRG individuals attending the gathering event on day t is calculated as

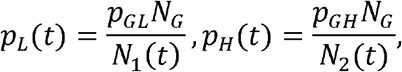

respectively, and the number of infectious attendees during the gathering at day t is given by

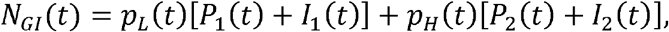

where 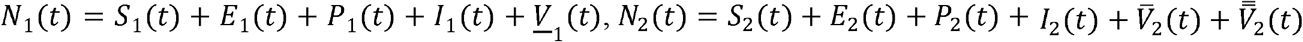. Note that the number of recovered individuals is small and not included for simplicity.

Following Champredon et al^20^, we calculate the expected minimum probability of transmissions per attendee that will occur during the gathering, considering the vaccination of attendees, yielding

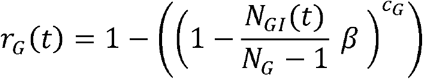

where *C*_*G*_ is the number of contacts with an infectious individual during the gathering. Hence, the number of LRG susceptible individuals infected during the gathering is

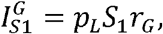

and the number of fully vaccinated LRG individuals infected during the gathering is

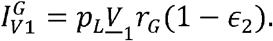

Similarly, we obtain the number of susceptible, the partially vaccinated and fully vaccinated individuals from HRG infected during the gathering as

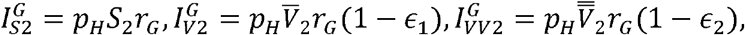

respectively.

The expected minimum number of transmissions that will occur during the gathering is thus

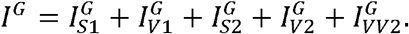

#### Intervention at gathering event

If the public health interventions, including vaccination strategy, are applied at the gathering event to prevent the transmission, we assume that only the fully vaccinated individuals are allowed to attend the gathering, which includes both vaccinated individuals and the individuals vaccinated but not effectively protected (infected, and in prodromal state). Hence, the proportion of attendance in the qualified LRG and HRG on day t is

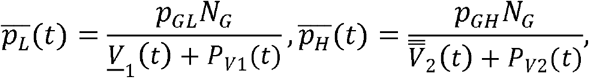

where 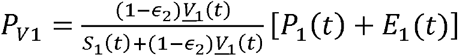 and 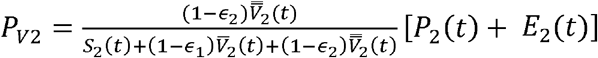 are the infection in the prodromal state in LRG and HRG and fully vaccinated individuals but not effectively protected, respectively.

Hence, the number of infectious attendees during the gathering on day t, which can only be the fully vaccinated individuals infected and without symptoms, is

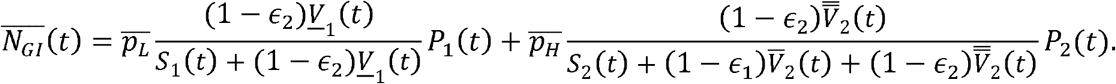

We also assume that only the fully vaccinated individuals are allowed to attend the gathering, hence the minimum probability of transmissions per attendee that will occur during the gathering becomes

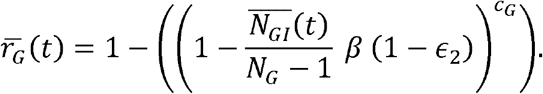

Hence, the expected minimum number of transmissions that will occur during the gathering is

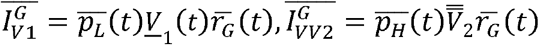

respectively.

### 2.6 Scenario analysis

We conduct numerical simulations with the setting of the hypothetical metropolitan city, starting on May 1, 2022, and run the model for 2 years (730 days). The vaccination started to be administered in HRG individuals on June 10, 2022^**Error! Reference source not found**.^. The initial values, parameters with fixed values, and range of some parameters used for simulations are presented in Tables 2 and 3. We investigate five different scenarios listed in Table 4, by presenting the projection of daily new infections in LRG and HRG (per 100 000 individuals). In scenarios 1-3, the projection of mean, and 95% confidence interval of daily new infections are obtained from 5000 parameter sets sampling from the prior distribution (uniform) of parameters, by the Latin hypercube sampling (LHS) method^30,31^. We only present the mean of all the simulations in scenarios 2 and 3 for an intuitive interpretation of results. We explore the proportion of vaccination coverage needed to prevent the transmission under different assumptions in scenarios 4 and 5, where other parameters are fixed at the values presented in Table 3. The analyses are conducted using MATLAB (R2020a)^32^.

**Table 2.**
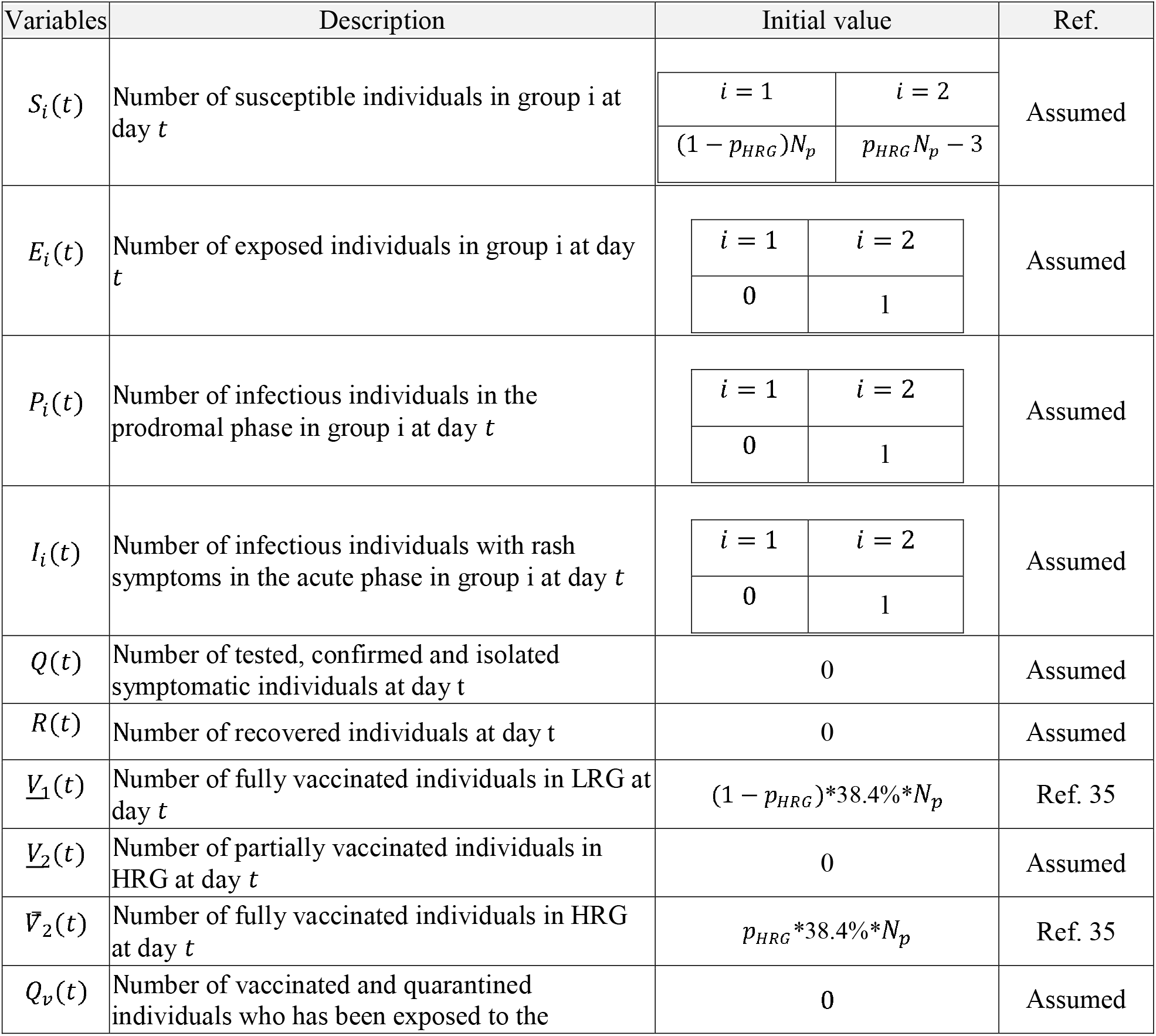
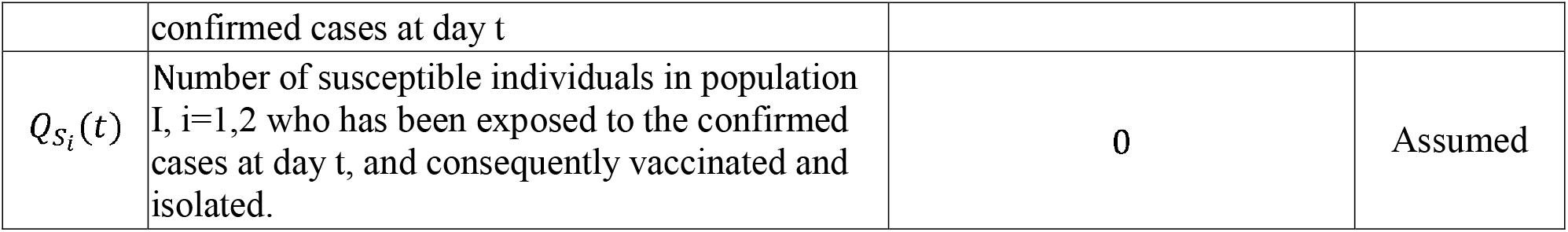
Variables used in the modelling of monkeypox transmission and their assumed initial values.

**Table 3.**
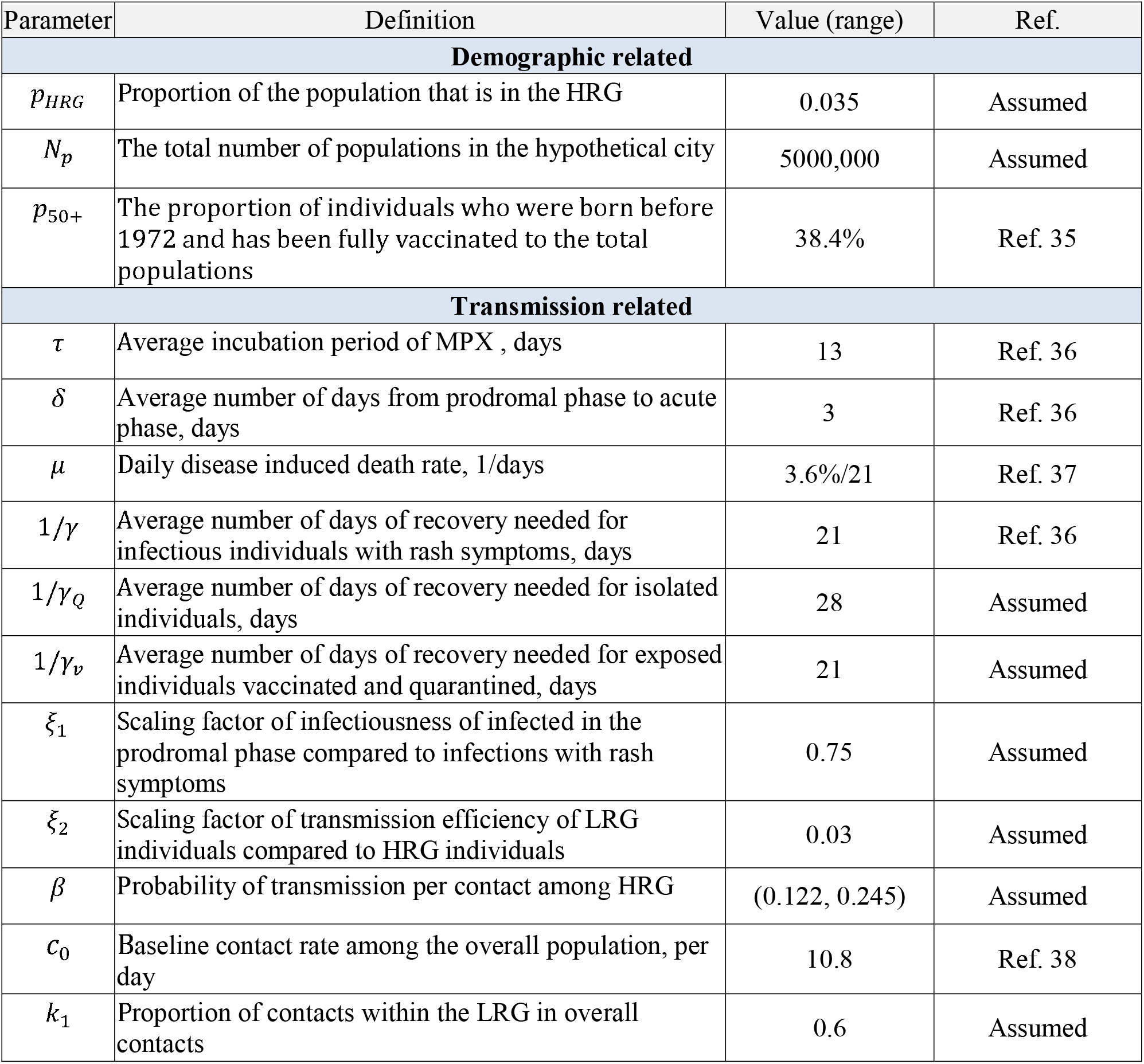
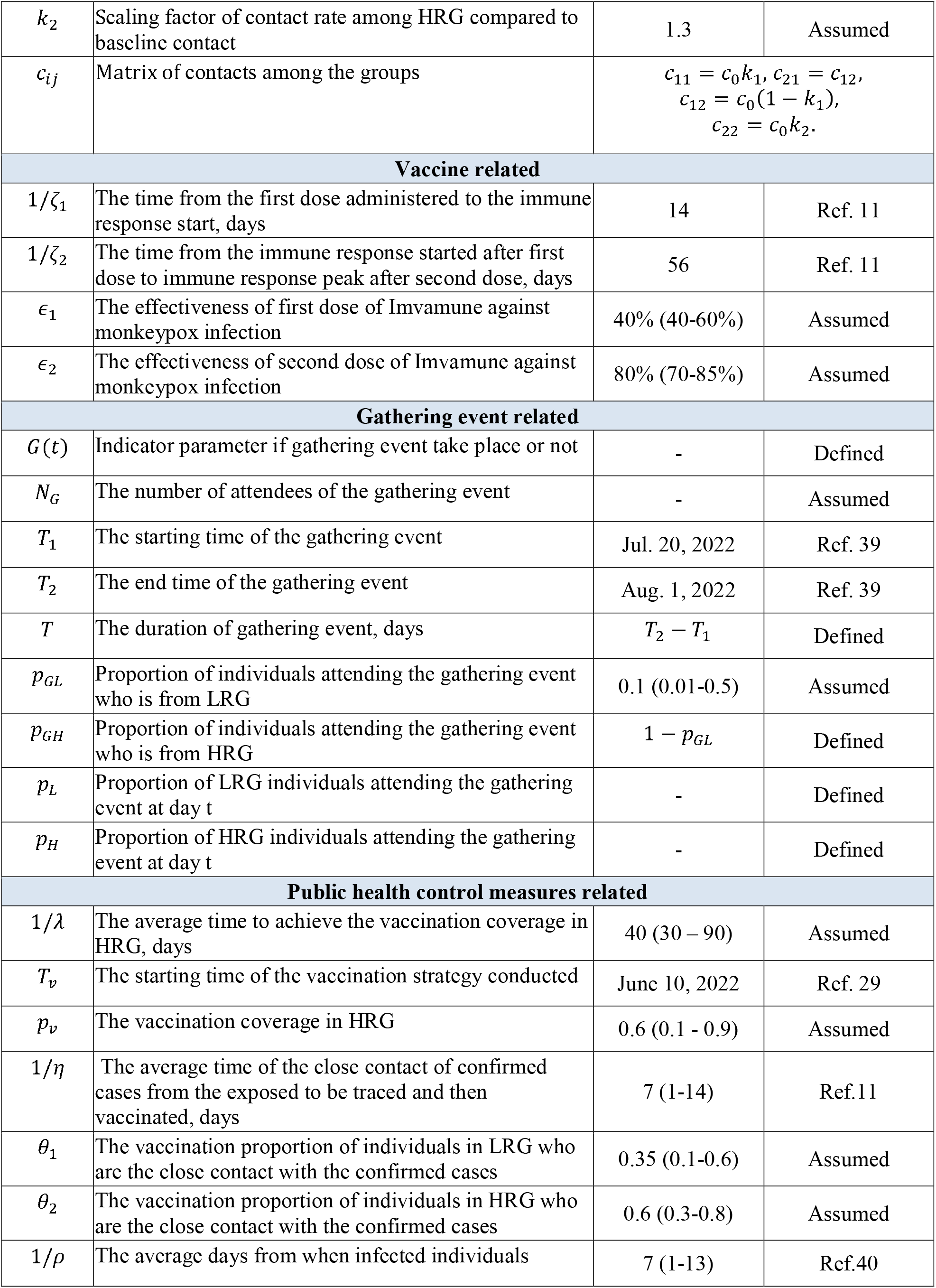
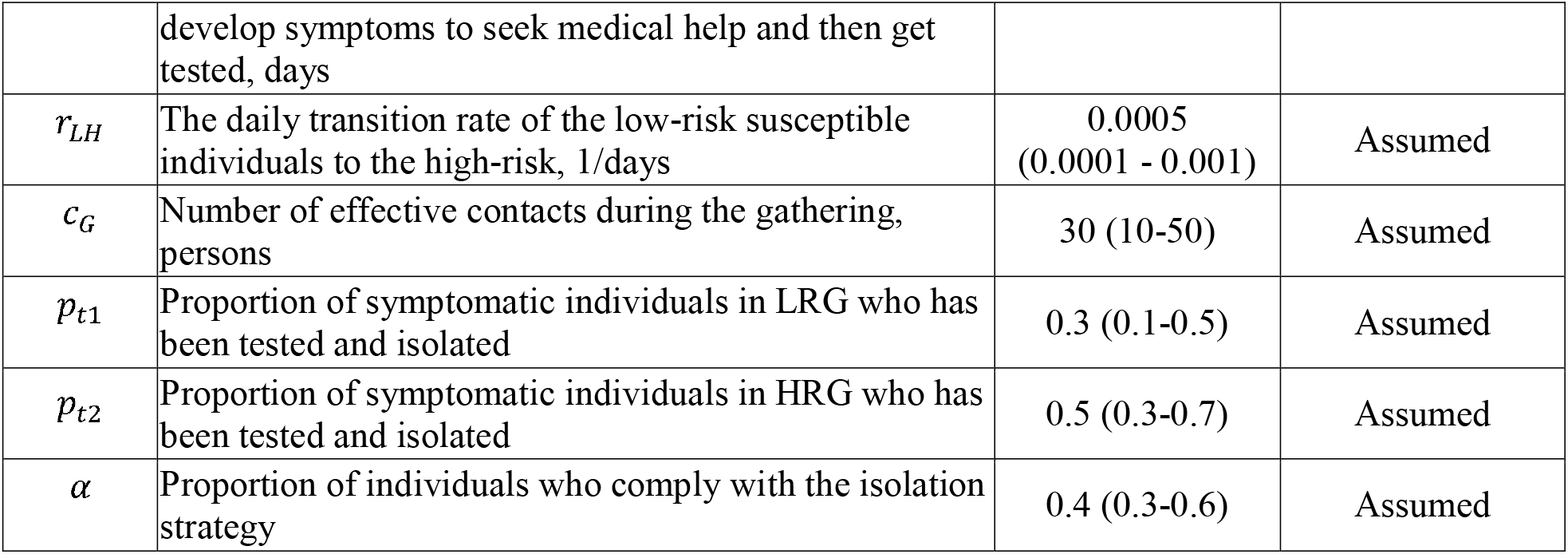
Parameters used in the modelling of monkeypox transmission.

**Table4.**
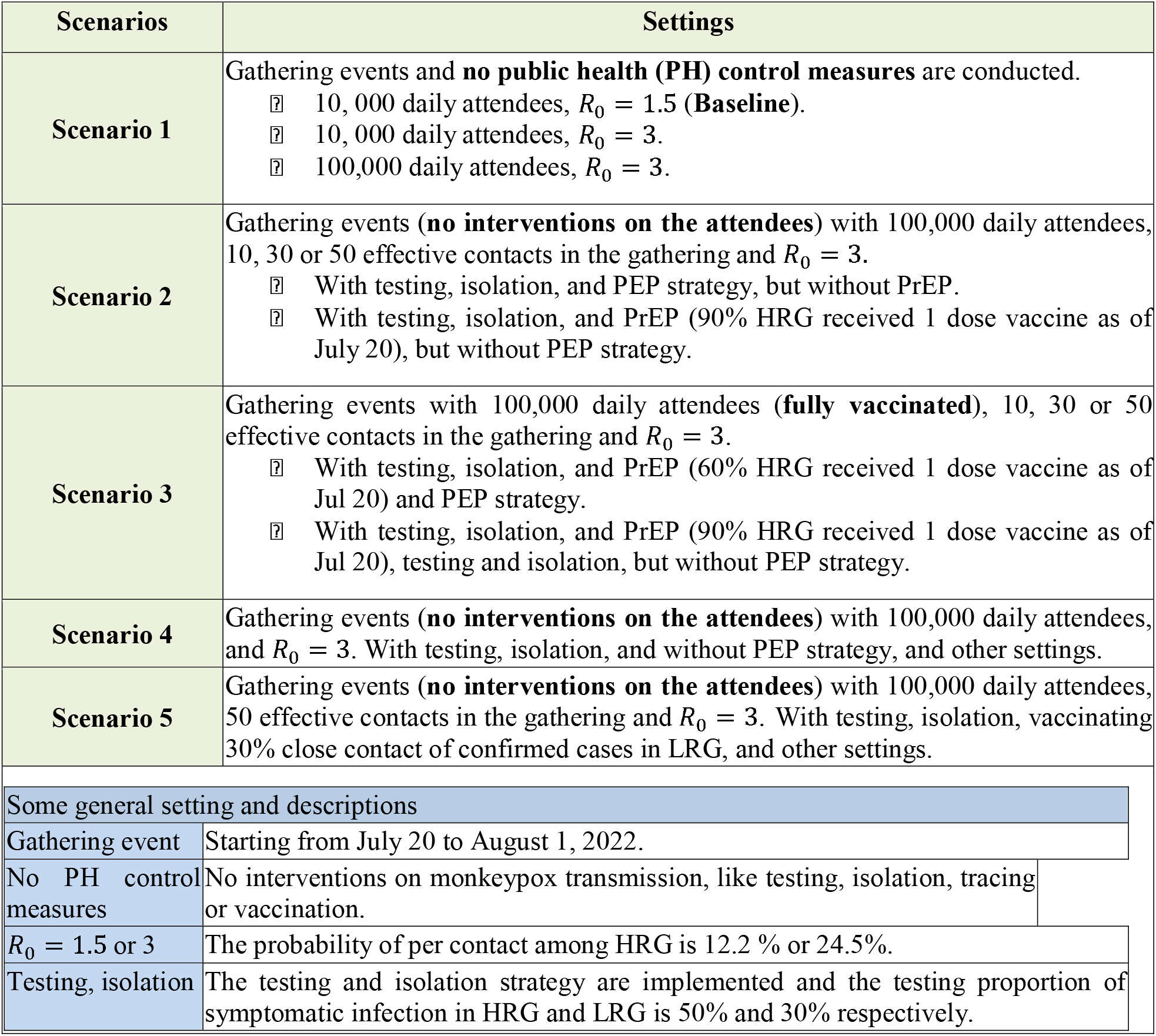
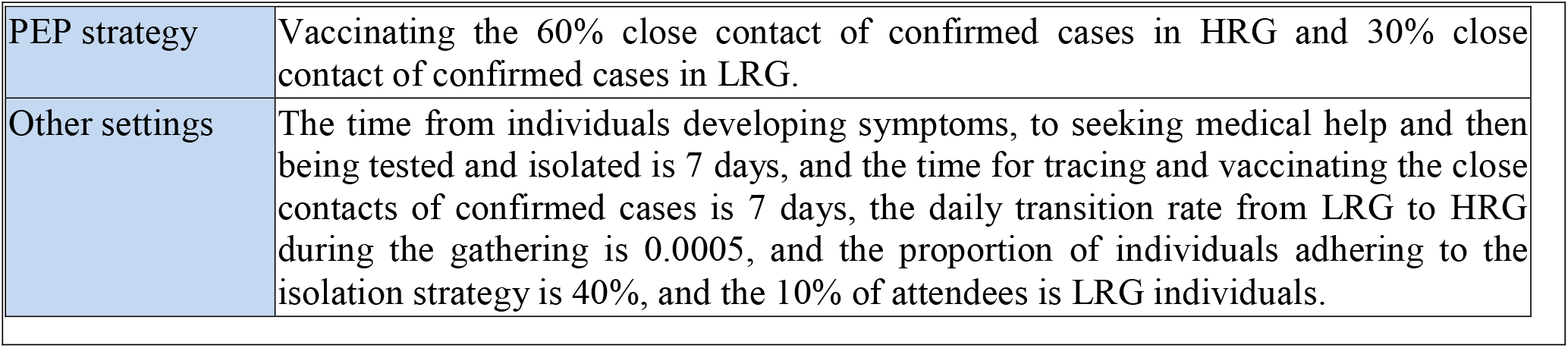
Lists of settings of scenarios to projections of monkeypox infection.

### 2.7 Sensitivity analysis

Sensitivity analyses are conducted to address the parameters’ uncertainty through the LHS and partial rank correlation coefficient (PRCC) method^30^. We generate 5000 samples of parameters related to vaccination strategy, including the efficacy of vaccine, the vaccination coverage in HRG, the days needed to achieve the vaccination coverage, and the vaccination proportion of close contacts of confirmed cases in HRG and LRG, as well as the parameters associated with gathering events, including the number of attendees of gathering, the effective contacts in the gathering, the proportion of attendees from LRG individuals. The ranges of parameters used in the sensitivity analysis are reported in Table A1 of the Appendix. We calculate the value of PRCC to investigate the relationship between the parameters and the model outputs of cumulative cases, which above 0.5 are considered to be significant.

## 3. Results

### 3.1 Impact of no public health measures implementation after gatherings

We project the daily new infections in LRG and HRG when the mass gathering events occur between July 20 to August 1 (**Figure 2**). We vary the number of participants and transmissibility levels of monkeypox virus, under the assumption that public health control measures are not in place. A large outbreak of cases follows mass gathering events as the number of attendees and transmissibility increase, raising the transmission. However, with the lowest number of participants and value of *R*_0_, the outbreak shows the beginning of an increasing trend 400 days after the gathering. For a gathering with 10,000 daily attendees, the risk of a monkeypox outbreak is low if the transmission probability per contact among the HRG individuals of the monkeypox virus is 12.2% (*R*_0_ = 1.5). On the other hand, we could observe a large outbreak if the number of participants increases to 100,000, with an average peak of daily new infections in HRG of around 150 per 100,000 people. If attendance increases to 100,000, the outbreak will immediately follow the gathering event, with a larger peak size of infections (500 per 100,000).

**Figure 2:**
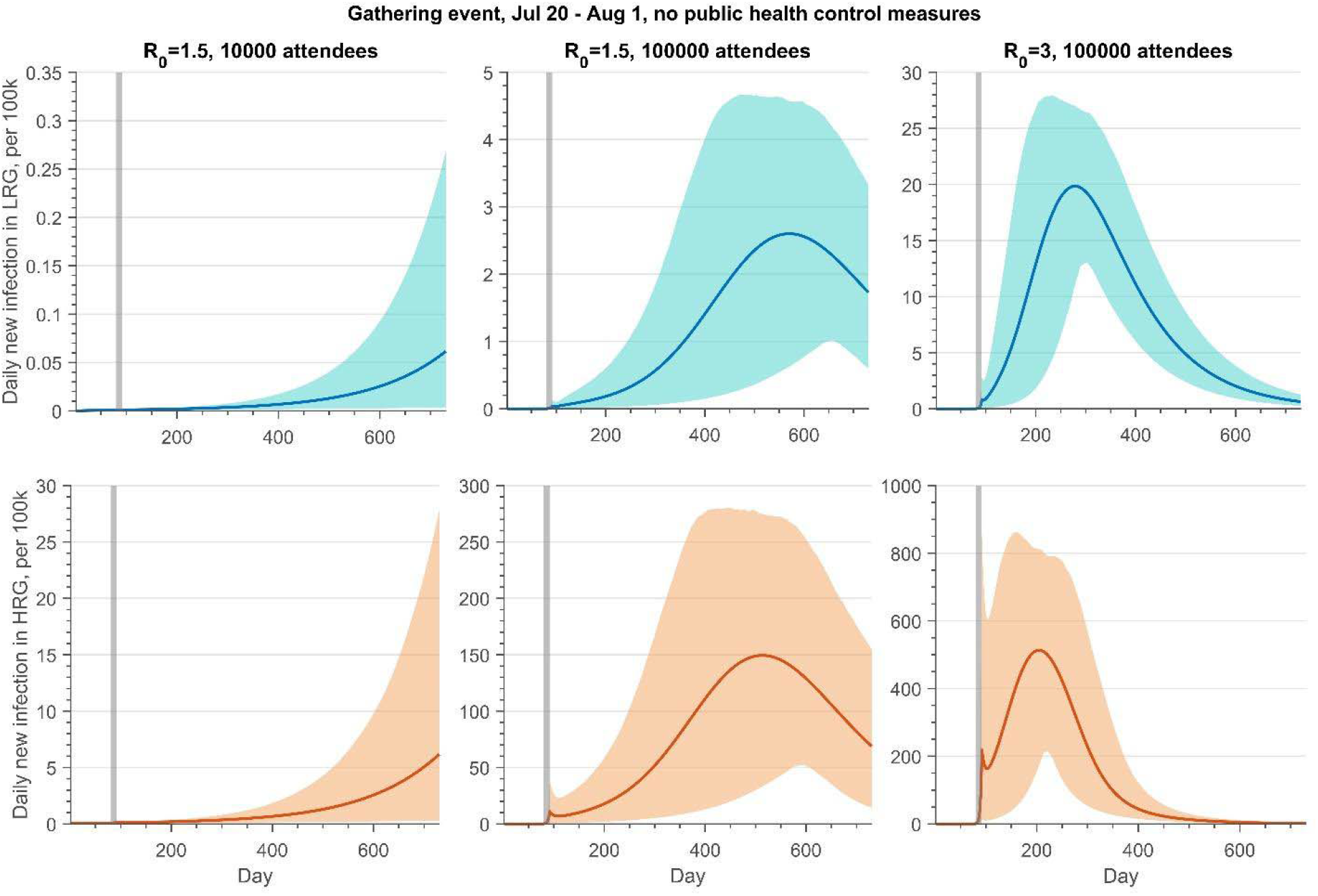
Projections of daily new infections (per 100k) in the LRG and HRG without public health control measures if there is a gathering event from Jul 20 to Aug 1, 2022, with 10000 or 100000 daily attendees. The reproduction number of monkeypox transmission is 1.5 or 3.

### 3.2 Benefits of PEP strategies

**Figures 3 and 4** show the impact of PEP and PrEP strategies under the possibility of a mass gathering with 100,000 attendees and relatively high transmission efficiency of the monkeypox virus (*R*_0_ = 3). Overall, if testing and isolation of symptomatic infection are in place, the PEP strategy is more beneficial to the control of monkeypox outbreaks, compared to the PrEP strategy, and this is expected given the time needed to develop an immune response. With the PEP strategy, vaccinating 60% and 30% of close contacts of confirmed monkeypox cases in HRG and LRG respectively, and maximum effective contacts in the gathering, the average peak of daily new infection in HRG was below 4 per 100,000 attendees (**Figure 3, second panel)**. However, this number exceeds 90 per 100,000 if 90% of HRG individuals received at least 1 dose vaccine before the gathering event started, but without PEP (**Figure 3, fourth panel)**.

**Figure 3:**
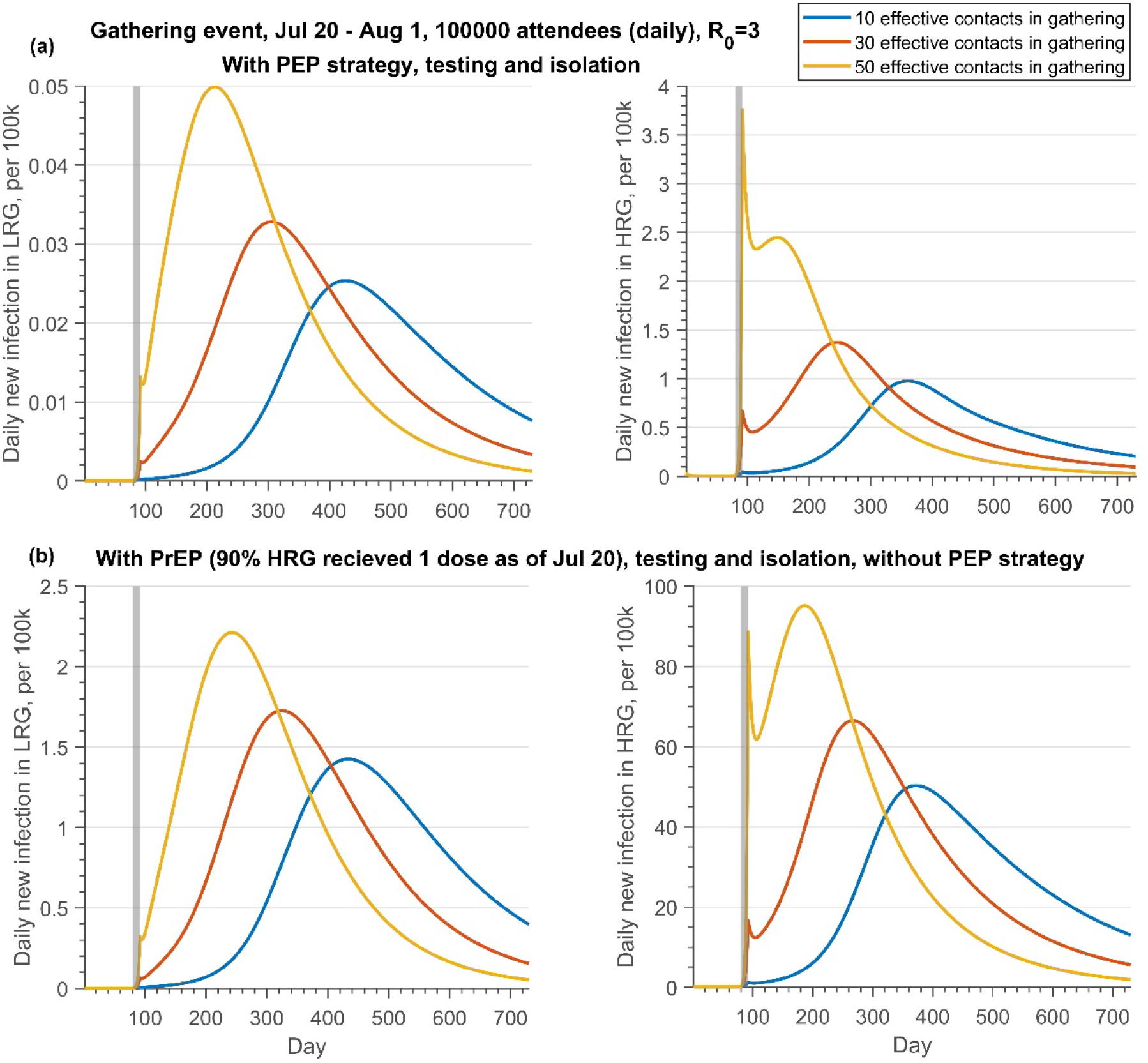
Projections of daily new infections (per 100k) in the LRG and HRG with different public health control strategies under the different effective contacts in the gathering if there is a gathering event occurring from Jul 20 to Aug 1, 2022, with daily 100000 attendees. (a) with PEP, testing and isolation, but without PrEP; (b) with PrEP, testing and isolation, but without PEP. Note that the reproduction number of monkeypox transmission is 3. The testing proportion of symptomatic infection in HRG and LRG is 50% and 30% respectively. The PEP represents that we vaccinated the 60% close contacts in HRG and 30% close contact in LRG. The grey shaded bar represents the gathering event occurring.

**Figure 4:**
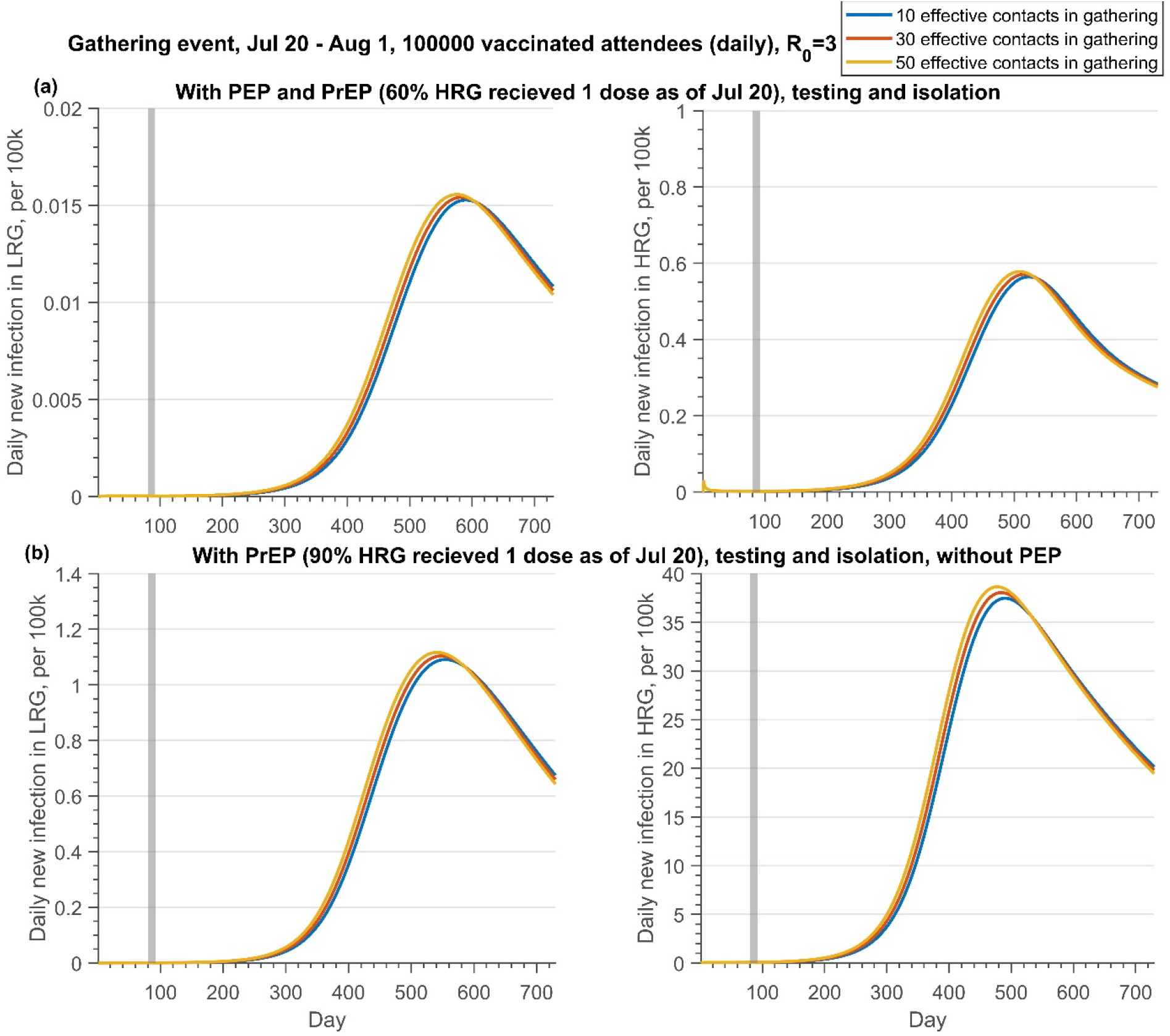
Projections of daily new infections (per 100k) in the LRG and HRG with different public health control strategies under the different effective contacts in the gathering if there is a gathering event occurring from Jul 20 to Aug 1, 2022, with daily 100000 attendees who has been fully vaccinated. (a) with PrEP (60% HRG received 1 dose before Jul 20) and PEP, testing and isolation; (b) with PrEP (90% HRG received 1 dose before Jul 20), testing and isolation, but without PEP. Note that the reproduction number of monkeypox transmission is 3. The testing proportion of symptomatic infection in HRG and LRG is 50% and 30% respectively. The PEP represents that we vaccinated the 60% close contacts in HRG and 30% close contact in LRG. The grey shaded bar represents the gathering event occurring.

Similar results are obtained if all attendees of the gathering event are fully vaccinated (**Figure 4**). If PEP strategy is implemented and the effective contacts during the gathering are 50, the mean peak of daily infection in HRG is below 1 per 100,000, although only 60% of individuals were administered 1 dose of vaccine before the gathering. Conversely, with only 10 effective contacts in the gathering, the mean peak of daily new infection in HRG exceeds 35 per 100, 000, if vaccinating 90% of HRG individuals with 1 dose before the gathering but without a PEP strategy. Regardless of whether there was a vaccination requirement on the attendance of gathering events, PEP strategies are critical to containing monkeypox transmission arising from gatherings.

Results for LRG follow the same trends of HRG, but with a smaller magnitude.

### 3.3 Measures to prevent outbreaks after gatherings: contact restriction or vaccination

The peak size of the monkeypox outbreak is significantly associated with the number of effective contacts in the gathering, if there are no restrictions on the gathering activities (**Figure 3**). Thus, the public health control measures aiming at constraining contact during the gathering are essential to prevent the possible outbreak under this circumstance.

However, the contacts during the gathering would have a slight effect on the progression of the monkeypox transmission if only fully vaccinated individuals are allowed to attend the gathering (**Figure 4**). Moreover, whether there is a vaccination requirement for the attendees of gathering activities or not, public health control measures, such as PEP, PrEP, testing, and isolation, are crucial to prevent the outbreak resulting from mass gathering events.

### 3.4 Identification of the best combination of vaccination coverage and gathering intervention

Figure 5. shows a contour plot of peak infection in LRG and HRG under **Scenario 4** (described in **Table 4**), when effective contacts and vaccine coverage (one dose by July 20) are varied. These results permit the determination of the vaccination coverage in HRG and constraints of contacts in gathering needed to prevent monkeypox transmission if the gathering event occurs. In this scenario, testing and isolation (30% and 50% of symptomatic infections in LRG and HRG) are included. As illustrated in **Figure 5**, public health interventions on constraining contacts during the gathering benefit more from containing the transmission, compared to the vaccination coverage in HRG. The infection in the LRG could be kept below 1 per 100,000 if the effective contacts in the gathering is 46. However, the contacts should be less than 15 to maintain a low prevalence (10 per 100,000) in HRG.

### 3.5 Identification of the best combination of PEP and PrEP strategy

The contour plot of peak infection in LRG and HRG with varying vaccination proportions of PEP and PrEP is shown in **Figure 6**, if all the individuals were allowed to attend the mass gathering events (100,000 attendees), along with assumptions listed in **Table 4 (Scenario 5)**. If vaccination coverage of 1 dose in HRG as of Jul 20 is 10%, to maintain the infection in LRG was below 1 per 100,000, at least 38% of close contact of confirmed cases in HRG should be vaccinated. However, this proportion needs to increase to 75% to keep the infection in HRG below 1 per 100,000.

**Figure 5:**
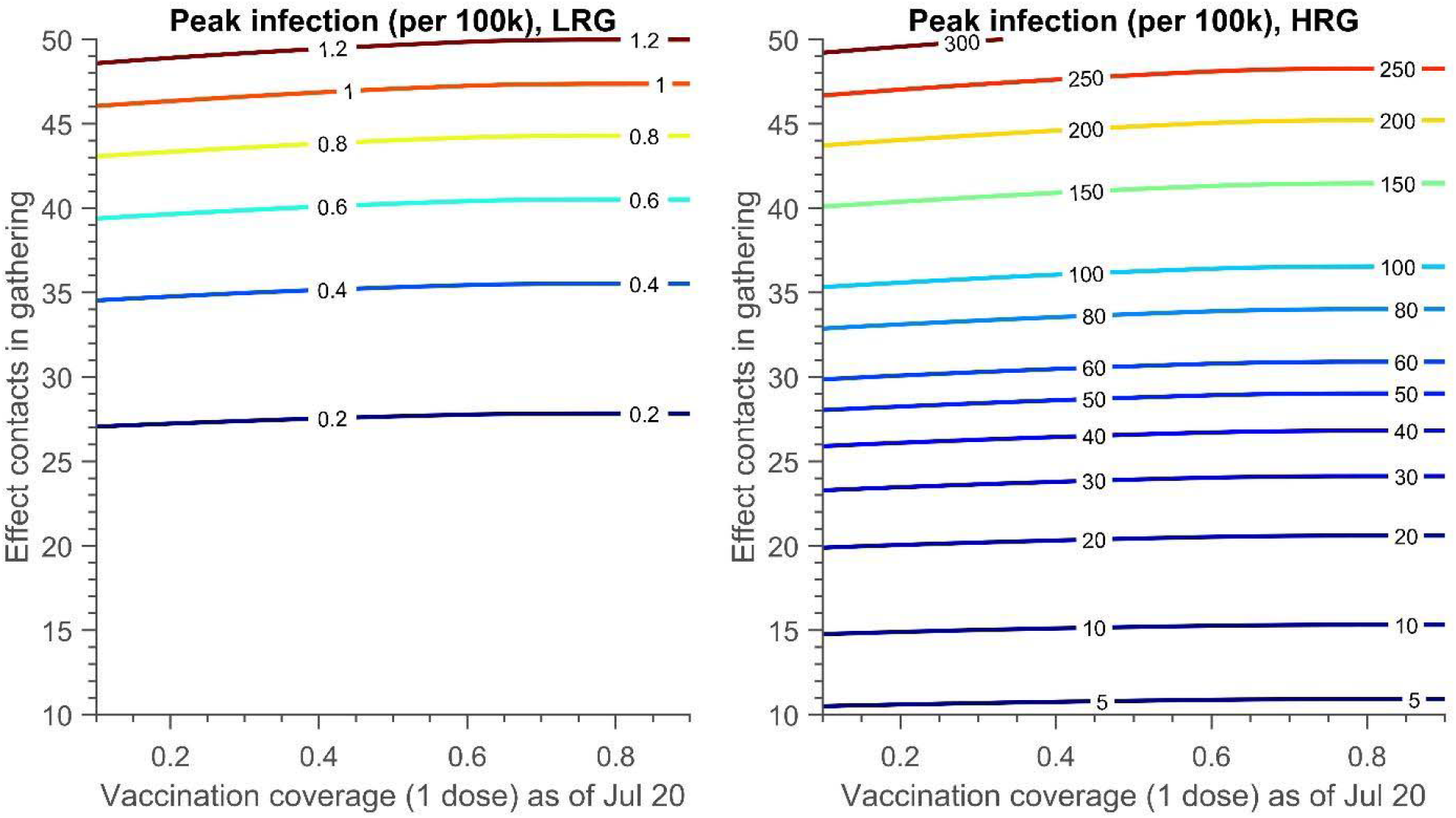
The contour plot of peak infection in LRG and HRG (per 100k) with different effect contacts in the gathering and the vaccination coverage achieved (1 dose) on Jul 20. There is no PEP strategy and the testing proportion of symptomatic infection in HRG and LRG is 50% and 30% respectively. The detailed setting can be found in Table 4, scenario 4.

**Figure 6:**
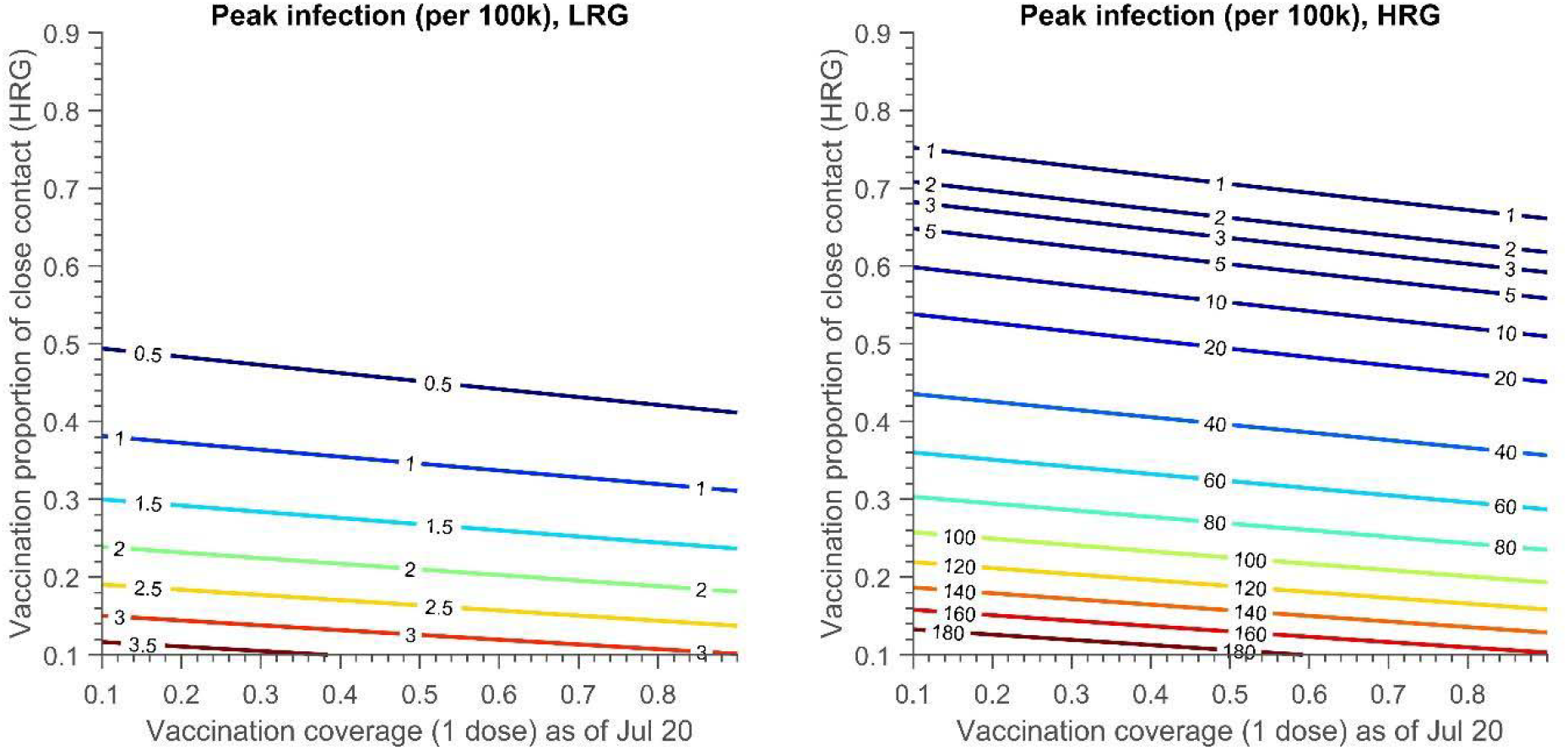
The contour plot of peak infection in LRG and HRG (per 100k) with vaccination proportion of close contact in HRG and the vaccination coverage achieved (1 dose) on Jul 20. The testing proportion of symptomatic infection in HRG and LRG is 50% and 30% respectively. 30% close contact of confirmed cases in LRG are traced, vaccinated and quarantined. The detailed setting can be found in Table 4, scenario 5.

### 3.6 Sensitivity analysis

The observed number of attendees, the number of effective contacts, and the transition rate from LRG to HRG during the gathering are significantly positively correlated with the cumulative cases (**Figure 7a**). On the other hand, the proportion of attendees from the LRG is negatively correlated to the cumulative cases, which indicates the dilution effect of the LRG attendees on the gathering events in consideration of the fact that they have a lower possibility to transmit the virus to the LRG community.

**Figure 7:**
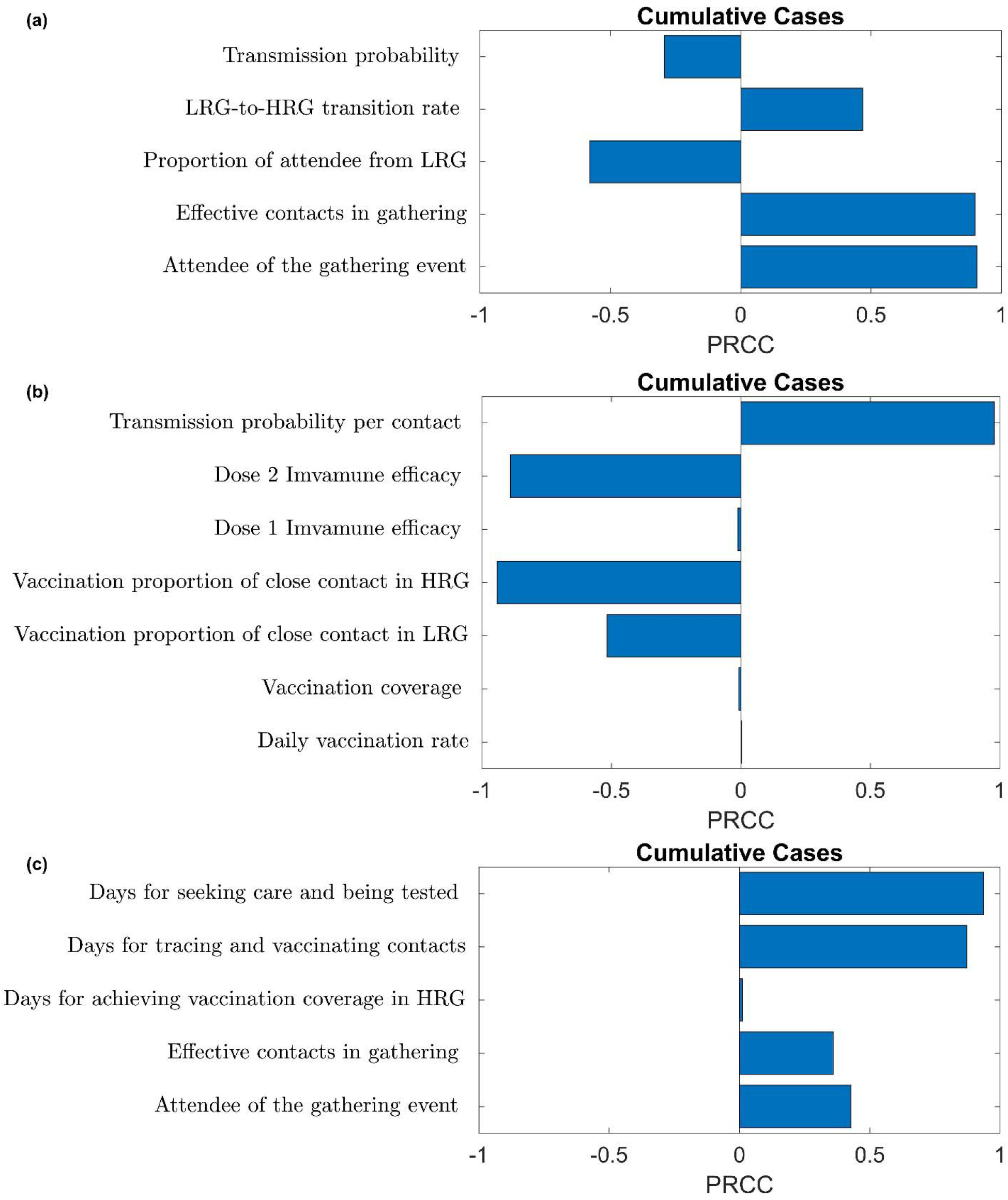
The PRCC plots of (a) gathering event-related (b) transmission and vaccine-related (c). public health control strategy-related parameters on cumulative cases.

The sensitivity analysis also shows that efficacy of the second dose, the vaccine coverage of close contact of confirmed cases in HRG and LRG present a negative correlation with cumulative cases (**Figure 7b**). Nevertheless, the first dose efficacy and its coverage in HRG, and the daily vaccination rate in HRG are not significant to the cumulative cases, due to the small proportion of HRG individuals in the total population (**Figure 7b**).

The sensitivity analysis confirms that the PEP strategy and the interventions in the gathering activities are crucial to curb monkeypox spreading at gatherings. Moreover, the efforts of contact tracing, testing and isolation of symptomatic monkeypox cases affect the progression of the disease significantly (**Figure 7c**).

## 4. Discussions

The highest alarm, PHEIC, on monkeypox by WHO signalled the seriousness of the global monkeypox outbreak^3^. In this study, we employ the mathematical modelling approach to explore how vaccination strategy can be implemented, in a hypothetical metropolitan city, to prevent the spreading of monkeypox in human populations after mass gatherings events. Our results suggest that the risk of a monkeypox outbreak after the gathering activities remains high, especially in the case the number of attendees is large and public health control measures are not in place. However, the effective vaccination strategy can support the containment of monkeypox transmission at mass gatherings where the constraints on effective contact in the gathering are carried out, along with the implementation of PrEP strategy and testing and isolation of symptomatic infection. On the other hand, the vaccination requirements for participants in mass gathering events also play a crucial role in curbing the spreading of viruses. In addition, tracing and vaccinating the close contacts who are exposed to the confirmed cases is more beneficial to contain monkeypox transmission compared to the targeted vaccination campaign in HRG individuals.

Our novel model structure, with consideration of saturation of contact at gatherings, allows us to assess the monkeypox transmission risk on the occasion that gathering activities occur. Our results deliver vital messages on how ring vaccination can be a powerful tool to halt the spread of the monkeypox virus linked to mass gathering events. Either limiting the gathering size and density, or requiring the vaccination of attendees, or both, is essential for safe social gathering events. Additional supports are also required, such as massive efforts on testing and isolation of confirmed cases and PEP strategy requesting rigorous contact tracing and the uptake of vaccination in close contacts of confirmed cases.

Nonetheless, the ultimate effect of the ring vaccination approach may not work as expected due to the uncertainty of efficacy and availability of stockpiles of the vaccine, as well as the difficulty in identifying the people who are most at risk from infection^32^. The effectiveness of smallpox vaccine against monkeypox infection in the human population remains unclear, although there are some positive evidences from animal studies. The rollout of the vaccine campaign in HRG, or extending the eligibility to individuals in middle risk, requires the support of enough stockpiles of the vaccine. Moreover, the difficulty to identify people with exposure to the virus should be taken into consideration, presuming that the PEP strategy is valued. Containing the spreading of monkeypox by ring vaccination protocol is greatly dependent on the willingness of inoculation of the individual with exposure and the compliance with the quarantine after vaccination. Targeting specific communities or groups of people may deepen stigma and hinder tracing, vaccination, and identification of cases ^6,33,34^.

Although our modelling scenario simulations are conducted in a hypothetical metropolitan city with parameters from literature with reference to Canadian cities, our model can be easily applied to any jurisdictions and areas where data are available, with the refinement of key parameters like the efficacy of vaccines to inform public health decision making. But the specific numbers required to halt the transmission need to be re-examined in a given region with local data. Besides, it is known that animals are a reservoir and part of the transmission to the human population, but this is not included in our work, and it can underestimate the transmission risk. We will include it in further study.

## Data Availability

All data produced in the present work are contained in the manuscript.

## Author Contributions

Conceptualization: H.Z., P.Y., Y.T., L.Y., E.A., N.O.; Data curation: Y.T., L.Y., P.Y.; Formal analysis, L.Y., P.Y., Y.T., H.Z.; Methodology: P.Y., Y.T., L.Y., E.A., H.Z.; Software: P.Y., Y.T.; Validation: P.Y., Y.T.; Visualization: P.Y., Y.T.; Writing - original draft: P.Y., Y.T., L.Y., E.A., H.Z.; Writing - review & editing: J.B., J.A., J.H., J.W., N.O., H.C., E.A., P.Y., Y.T., L.Y., H.Z..; Funding acquisition: H.Z., J.A., J.B., J.H., J.W., H.C.; Supervision: H.Z.

