## Appendix for "Modelling vaccination and control strategies of outbreaks of monkeypox at gatherings"

**Affiliations**

4700 Keele Street, Toronto, Ontario, Canada, M3J1P3

### A1. Model

#### Model equations

Following the flowchart in Figure 1, the transmission of monkeypox in the area is described by the following system of differential equations

Following the flowchart in Figure 1, the transmission of monkeypox in the area is described by the following system of differential equations

$$
\begin{cases}
S_1' = -F_1 S_1[1 - G(t)p_L] - G(t)I_{S1}^G - r_{LH}G(t)S_1 - \theta_1\eta Z_{S1}, \\
E_1' = F_1[S_1 + (1-\epsilon_2)\underline{V}_1][1 - G(t)p_L] + G(t)(I_{S1}^G + I_{V1}^G) - \dfrac{E_1 - \theta_1\eta Z_{E1}}{\tau} - \theta_1\eta Z_{E1}, \\
P_1' = \dfrac{E_1 - \theta_1\eta Z_{E1}}{\tau} - \dfrac{P_1 - \theta_1\eta Z_{P1}}{\delta} - \theta_1\eta Z_{P1}, \\
I_1' = \dfrac{P_1 - \theta_1\eta Z_{P1}}{\delta} - p_{t1}\alpha\rho I_1 - (1 - p_{t1}\alpha)\gamma I_1 - \mu I_1, \\
S_2' = -F_2 S_2[1 - G(t)p_H] - G(t)I_{S2}^G - \lambda\zeta_1 S_2 + r_{LH}G(t)S_1 - \theta_2\eta Z_{S2}, \\
E_2' = F_2\big[S_2 + (1-\epsilon_1)\bar{V}_2 + (1-\epsilon_2)\bar{\bar{V}}_2\big][1 - G(t)p_H] + G(t)(I_{S2}^G + I_{V2}^G + I_{VV2}^G) - \dfrac{E_2 - \theta_2\eta Z_{E2}}{\tau} - \theta_2\eta Z_{E2}, \\
P_2' = \dfrac{E_2 - \theta_2\eta Z_{E2}}{\tau} - \dfrac{P_2 - \theta_2\eta Z_{P2}}{\delta} - \theta_2\eta Z_{P2}, \\
I_2' = \dfrac{P_2 - \theta_2\eta Z_{P2}}{\delta} - p_{t2}\alpha\rho I_2 - (1 - p_{t2}\alpha)\gamma I_2 - \mu I_2, \\
Q' = p_{t1}\alpha\rho I_1 + p_{t2}\alpha\rho I_2 - \gamma_Q Q - \mu Q, \\
R' = (1 - p_{t1}\alpha)\gamma I_1 + (1 - p_{t2}\alpha)\gamma I_2 + \gamma_Q Q + \gamma_v Q_v, \\
\underline{V}_1' = -(1-\epsilon_2)F_1\underline{V}_1[1 - G(t)p_L] - G(t)I_{V1}^G + \zeta Q_{s1}, \\
\bar{V}_2' = \lambda\zeta_1 S_2 - (1-\epsilon_1)F_2\bar{V}_2[1 - G(t)p_H] - G(t)I_{V2}^G - \zeta_2\bar{V}_2, \\
\bar{\bar{V}}_2' = \zeta_2\bar{V}_2 - (1-\epsilon_2)F_2\bar{\bar{V}}_2[1 - G(t)p_H] - G(t)I_{VV2}^G + \zeta Q_{s2}, \\
Q_v' = \theta_1\eta Z_{E1} + \theta_1\eta Z_{P1} + \theta_2\eta Z_{E2} + \theta_2\eta Z_{P2} - \gamma_v Q_v, \\
Q_{s1}' = \theta_1\eta Z_{s1} - \zeta Q_{s1}, \\
Q_{s2}' = \theta_2\eta Z_{s2} - \zeta Q_{s2},
\end{cases}
$$

where $F_1 = \frac{\xi_2\beta}{N}\{c_{11}(\xi_1 P_1 + I_1)[1 - G(t)p_L] + c_{12}(\xi_1 P_2 + I_2)[1 - G(t)p_H]\}$,

$F_2 = \frac{\beta}{N}\{c_{21}(\xi_1 P_1 + I_1)[1 - G(t)p_L] + c_{22}(\xi_1 P_2 + I_2)[1 - G(t)p_H]\}$, $N = N_1 + N_2 + R$,

$G = \begin{cases} 1, & t \in [T_1, T_2], \\ 0, t < T_1, t > T_2. \end{cases}$, and $\lambda = \begin{cases} 0, & t < T_v, \\ \lambda, & t \geq T_v. \end{cases}$

**The model with intervention of gathering event**

$$
\begin{cases}
S_1' = -F_1S_1 - r_{LH}G(t)S_1 - \theta_1\eta Z_{S1}, \\
E_1' = F_1S_1 + F_1[(1-\epsilon_2)\underline{V_1}][1-G(t)\overline{p_L}] + G(t)\overline{I_{V1}^G} - \dfrac{E_1 - \theta_1\eta Z_{E1}}{\tau} - \theta_1\eta Z_{E1}, \\
P_1' = \dfrac{E_1 - \theta_1\eta Z_{E1}}{\tau} - \dfrac{P_1 - \theta_1\eta Z_{P1}}{\delta} - \theta_1\eta Z_{P1}, \\
I_1' = \dfrac{P_1 - \theta_1\eta Z_{P1}}{\delta} - p_{t1}\alpha\rho I_1 - (1-p_{t1}\alpha)\gamma I_1 - \mu I_1, \\
S_2' = -F_2S_2 - \lambda\zeta_1 S_2 + r_{LH}G(t)S_1 - \theta_2\eta Z_{S2}, \\
E_2' = F_2[S_2 + (1-\epsilon_1)\bar{V}_2] + F_2(1-\epsilon_2)\bar{\bar{V}}_2[1-G(t)\overline{p_H}] + G(t)\overline{I_{VV2}^G} - \dfrac{E_2 - \theta_2\eta Z_{E2}}{\tau} - \theta_2\eta Z_{E2}, \\
P_2' = \dfrac{E_2 - \theta_2\eta Z_{E2}}{\tau} - \dfrac{P_2 - \theta_2\eta Z_{P2}}{\delta} - \theta_2\eta Z_{P2}, \\
I_2' = \dfrac{P_2 - \theta_2\eta Z_{P2}}{\delta} - p_{t2}\alpha\rho I_2 - (1-p_{t2}\alpha)\gamma I_2 - \mu I_2, \\
Q' = p_{t1}\alpha\rho I_1 + p_{t2}\alpha\rho I_2 - \gamma_Q Q - \mu Q, \\
R' = (1-p_{t1}\alpha)\gamma I_1 + (1-p_{t2}\alpha)\gamma I_2 + \gamma_Q Q + \gamma_v Q_v, \\
\underline{V_1}' = -(1-\epsilon_2)F_1\underline{V_1}[1-G(t)\overline{p_L}] - G(t)\overline{I_{V1}^G} + \zeta Q_{s1}, \\
\bar{V}_2' = \lambda\zeta_1 S_2 - (1-\epsilon_1)F_2\bar{V}_2 - \zeta_2\bar{V}_2, \\
\bar{\bar{V}}_2' = \zeta_2\bar{V}_2 - (1-\epsilon_2)F_2\bar{\bar{V}}_2[1-G(t)\overline{p_H}] - G(t)\overline{I_{VV2}^G} + \zeta Q_{s2}, \\
Q_v' = \theta_1\eta Z_{E1} + \theta_1\eta Z_{P1} + \theta_2\eta Z_{E2} + \theta_2\eta Z_{P2} - \gamma_v Q_v, \\
Q_{s1}' = \theta_1\eta Z_{s1} - \zeta Q_{s1}, \\
Q_{s2}' = \theta_2\eta Z_{s2} - \zeta Q_{s2},
\end{cases}
$$

where $F_1 = \frac{\xi_2\beta}{N}\{c_{11}(\xi_1P_1 + I_1)[1-G(t)\overline{p_L}] + c_{12}(\xi_1P_2 + I_2)[1-G(t)\overline{p_H}]\}$,

$F_2 = \frac{\beta}{N}\{c_{21}(\xi_1P_1 + I_1)[1-G(t)p_L] + c_{22}(\xi_1P_2 + I_2)[1-G(t)p_H]\}$, $N = N_1 + N_2 + R$,

$G = \begin{cases} 1, & t \in [T_1, T_2], \\ 0, t < T_1, t > T_2. \end{cases}$, and $\lambda = \begin{cases} 0, & t < T_v, \\ \lambda, & t \geq T_v. \end{cases}$

**The simplified model without public health control measures**

$$\begin{cases} S_1' = -F_1 S_1, \\ E_1' = F_1 S_1 - \dfrac{E_1}{\tau}, \\ P_1' = \dfrac{E_1}{\tau} - \dfrac{P_1}{\delta}, \\ I_1' = \dfrac{P_1}{\delta} - \gamma I_1 - \mu I_1, \\ S_2' = -F_2 S_2, \\ E_2' = F_2 S_2 - \dfrac{E_2}{\tau}, \\ P_2' = \dfrac{E_2}{\tau} - \dfrac{P_2}{\delta}, \\ I_2' = \dfrac{P_2}{\delta} - \alpha\rho I_2 - (1-\alpha)\gamma I_2 - \mu I_2, \\ R' = \gamma I_1 + \gamma I_2, \end{cases}$$

where $F_1 = \frac{\xi_2 \beta}{N}\{c_{11}(\xi_1 P_1 + I_1) + c_{12}(\xi_1 P_2 + I_2)\}$, $F_2 = \frac{\beta}{N}\{c_{21}(\xi_1 P_1 + I_1) + c_{22}(\xi_1 P_2 + I_2)\}$, $N = N_1 + N_2 + R$.

### A2. The basic reproduction number

For the simplified model without public health control measures, we calculated the basic reproduction number following the next generation matrix method, yielding

$$R_0 = \frac{R_{01} + R_{02}}{2} + \frac{\sqrt{(R_{01} - R_{02})^2 + 4R_{012}R_{021}}}{2},$$

$$R_{01} = (1 - p_{hrg})\beta\xi_2 c_{11}\left[\frac{1}{\gamma + \mu} + \delta\xi_1\right], R_{02} = p_{hrg}\beta c_{22}\left[\frac{1}{\gamma + \mu} + \delta\xi_1\right],$$

$$R_{012} = (1 - p_{hrg})\beta\xi_2 c_{12}\left[\frac{1}{\gamma + \mu} + \delta\xi_1\right], R_{021} = p_{hrg}\beta c_{21}\left[\frac{1}{\gamma + \mu} + \delta\xi_1\right].$$

We fixed other parameters presented in Table 3 and then presented the contour plot of reproduction numbers without public health control measures under different transmission probability per contact among HRG and the proportion of the HRG individuals to the total populations in **Figure A1**. The transmission risk of the monkeypox virus in a city may increase with the proportion of the HRG individuals to the total population and if this number reaches a certain level, there is a spillover effect from HRG to LRG (**Figure A1b**).

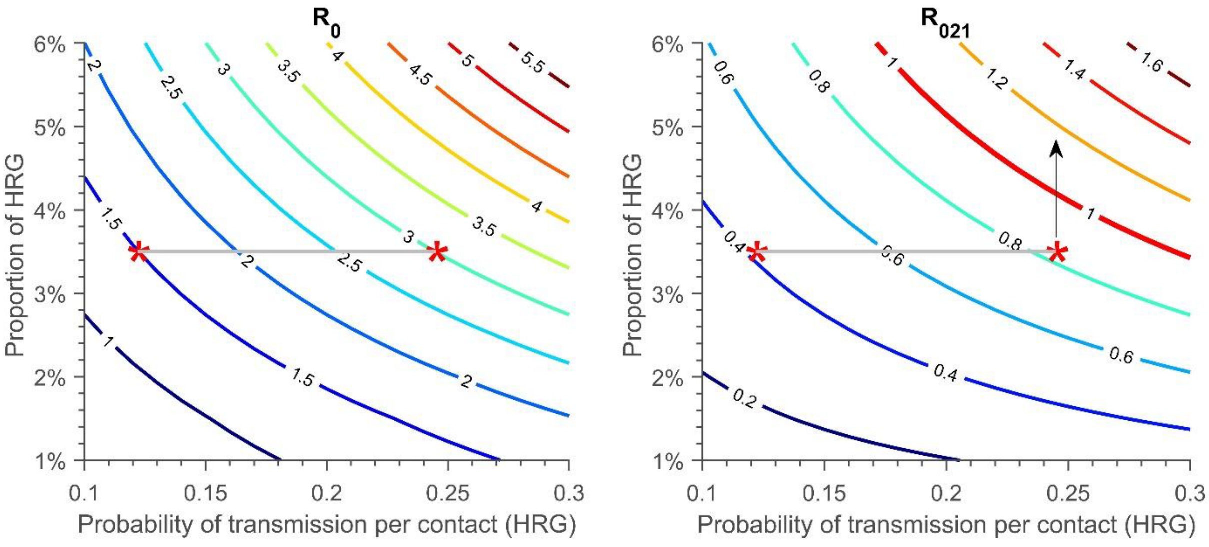


(a) (b)

**Figure A1:** The contour plot of reproduction number without public health control measures: (a) total population and (b) from HRG to LRG population with different probability of transmission per contact among HRG and the proportion of HRG in the total population. The red stars show the parameters used in our simulations and the grey lines indicate the range of parameters.

#### A3. Sensitivity analysis

**Table A1:** Parameters range for the Latin Hypercube Sampling (LHS) in the sensitivity analysis

| Parameter | | Definition | Range |
|---|---|---|---|
| Gathering related | $N_G$ | The number of attendees of the gathering event | $[5000, 50000]$ |
| | $c_G$ | Number of effective contacts during the gathering, persons | $[10, 50]$ |
| | $p_{GL}$ | Proportion of individuals attending the gathering event who is from LRG | $[0.01, 0.2]$ |
| | $r_{LH}$ | The daily transition rate of the low-risk susceptible individuals to the high-risk, 1/days | $[0.0001, 0.001]$ |
| Vaccine related | $1/\lambda_v$ | Average number of days of recovery needed for exposed individuals vaccinated and quarantined, days | $[0.001, 0.01]$ |
| | $p_v$ | The vaccination coverage in HRG | $[0.01, 1]$ |
| | $\theta_1$ | The vaccination proportion of individuals in LRG who are the close contact with the confirmed cases | $[0.1, 0.8]$ |
| | $\theta_2$ | The vaccination proportion of individuals in HRG who are the close contact with the confirmed cases | $[0.1, 1]$ |
| | $\epsilon_1$ | The effectiveness of first dose of Imvamune against monkeypox infection | $[0.01, 1]$ |
| | $\epsilon_2$ | The effectiveness of second dose of Imvamune against monkeypox infection | $[0.01, 1]$ |
| Testing and tracing related | $1/\lambda$ | The average time to achieve the vaccination coverage in HRG, days | [30, 90] |
| | $1/\eta$ | The average time of the close contact of confirmed cases from the exposed to be traced and then vaccinated, days | [1, 14] |
| | $1/\rho$ | The average days from when infected individuals develop symptoms to seek medical help and then get tested, days | [1, 13] |
